# Estimating the number of COVID-19-related infections, deaths and hospitalizations in Iran under different physical distancing and isolation scenarios: A compartmental mathematical modeling

**DOI:** 10.1101/2020.04.22.20075440

**Authors:** Hamid Sharifi, Yunes Jahani, Ali Mirzazadeh, Milad Ahmadi Gohari, Mehran Nakhaeizadeh, Mostafa Shokoohi, Sana Eybpoosh, Hamid Reza Tohidinik, Ehsan Mostafavi, Davood Khalili, Seyed Saeed Hashemi Nazari, Mohammad Karamouzian, Ali Akbar Haghdoost

## Abstract

**Background:** Iran is one of the countries that has been overwhelmed with COVID-19. We aimed to estimate the total number of COVID-19 related infections, deaths, and hospitalizations in Iran under different physical distancing and isolation scenarios.

**Methods:** We developed a Susceptible-Exposed-Infected-Removed (SEIR) model, parameterized to the COVID-19 pandemic in Iran. We used the model to quantify the magnitude of the outbreak in Iran and assess the effectiveness of isolation and physical distancing under five different scenarios (A: 0% isolation, through E: 40% isolation of all infected cases). We used Monte-Carlo simulation to calculate the 95% uncertainty intervals (UI).

**Findings:** Under scenario A, we estimated 5,196,000 (UI 1,753,000 - 10,220,000) infections to happen till mid-June with 966,000 (UI 467,800 - 1,702,000) hospitalizations and 111,000 (UI 53,400 - 200,000) deaths. Successful implantation of scenario E would reduce the number of infections by 90% (i.e. 550,000) and change the epidemic peak from 66,000 on June 9^th^ to 9,400 on March 1^st^. Scenario E also reduces the hospitalizations by 92% (i.e. 74,500), and deaths by 93% (i.e. 7,800).

**Interpretation:** With no approved vaccination or therapy, we found physical distancing and isolation that includes public awareness and case-finding/isolation of 40% of infected people can reduce the burden of COVID-19 in Iran by 90% by mid-June.

**Funding:** We received no funding for this work.

**Research in context:** *Evidence before this study:* Iran has been heavily impacted by the COVID-19 outbreak, and the virus has now spread to all of its provinces. Iran has been implementing different levels of partial physical distancing and isolation policies in the past few months. We searched PubMed and preprint archives for articles published up to April 15, 2020 that included information about control measures against COVID-19 in Iran using the following terms: (“coronavirus” OR “2019-nCoV” OR “COVID-19”) AND “Iran” AND (“intervention” OR “prevention” OR “physical distancing” OR “social distancing”). We found no studies that had quantified the impact of policies in Iran.

*Added value of this study:* Given the scarcity of evidence on the magnitude of the outbreak and the burden of COVID-19 in Iran, we used multiple sources of data to estimate the number of COVID-19 infections, hospitalizations, and deaths under different physical distancing and isolation scenarios until mid-June. We showed that implementing no control measures could lead to over five million infections in Iran; ∼19% of whom would be hospitalized, and ∼2% would die. However, under our most optimistic scenario, these estimates could be reduced by ∼90%.

*Implications of all the available evidence:* With no effective vaccination or treatment, advocating and enforcing physical distancing and isolation along with public education on prevention measures could significantly reduce the burden of COVID-19 in Iran. Nonetheless, even under the most optimistic scenario, the burden of COVID-19 would be substantial and well beyond the current capacity of the healthcare system in Iran.

## Introduction

The COVID-19 was declared a pandemic on March 11, 2020 and the disease is now expected to spread worldwide. The risk is relatively low for the general population, although people aged 65 years and over, those with suppressed immune systems, and people with underlying medical conditions (e.g., cardiovascular or respiratory diseases) are at increased risk of adverse outcomes. The case fatality rate of the infection is estimated to be around 2% (95% CI: 2-3%) and as of April 15, 2020, a total number of 2,049,888 confirmed cases, 510,486 recovered cases, and 133,572 death have been reported worldwide.^1^ Iran is one of the hardest hit countries by COVID-19 and has been struggling with controlling the disease for over two months. The first confirmed cases of COVID-19-related deaths were reported on January 21 in the city of Qom; 200 Km away from Iran’s capital city of Tehran. As of April 15, 2020, a total number of 76,389 confirmed cases, 49,933 recovered cases, and 4,777 death have been reported and COVID-19 has spread to all of its provinces;^2^ figures that are highest among the Eastern Mediterranean Region countries.^3^

The Susceptible-Exposed-Infected-Removed (SEIR) model provides a mathematical framework to explain the spread of infectious diseases and has previously been used for estimating the epidemiological parameters of several infectious diseases such as measles, Ebola, and influenza.^4–6^ SEIR could also help evaluate the impact of implementing various interventions (e.g., isolation and physical distancing policies) aimed at controlling the growth of the pandemic and flattening the epidemic curve. Physical distancing (also called social distancing) control measures are policies that aim to minimize close contacts within communities and include individual-level (e.g., quarantine, isolation) and community-level (closure of educational and recreational settings, non-essential businesses, and cancellation of public/mass/crowded gatherings) approaches.^7,8^

In Iran, the physical distancing and isolation interventions were scaled up in late February and early March by nationwide closure of schools/universities, cancellation of sports events and Friday/congregational prayers as well as the closure of all non-essential services, tourism sites, and shopping malls (Figure 1). Iran also closed its holy shrines in Mashhad and Qom in mid-March. Moreover, while there were no mandatory shelter-in-place or lockdown orders, people were encouraged to stay at home. People were also asked to avoid travelling during the New year Holidays (i.e., Nowruz) from March 19 to 26; however, no restrictions for domestic (via flight, train or bus) or international travels (i.e. no border closure) were imposed.^9^ Despite implementing various physical distancing control measures, our understanding of their impact on the magnitude of COVID-19-related new infections, hospitalization, and death remains limited. In this study, we aim to provide an estimate of these epidemiological parameters and approximate the peak date of the epidemic in Iran under different physical distancing and isolation scenarios. These estimates are of particular importance for COVID-19-related health policy, planning, and financing purposes in Iran.

**Figure 1.**
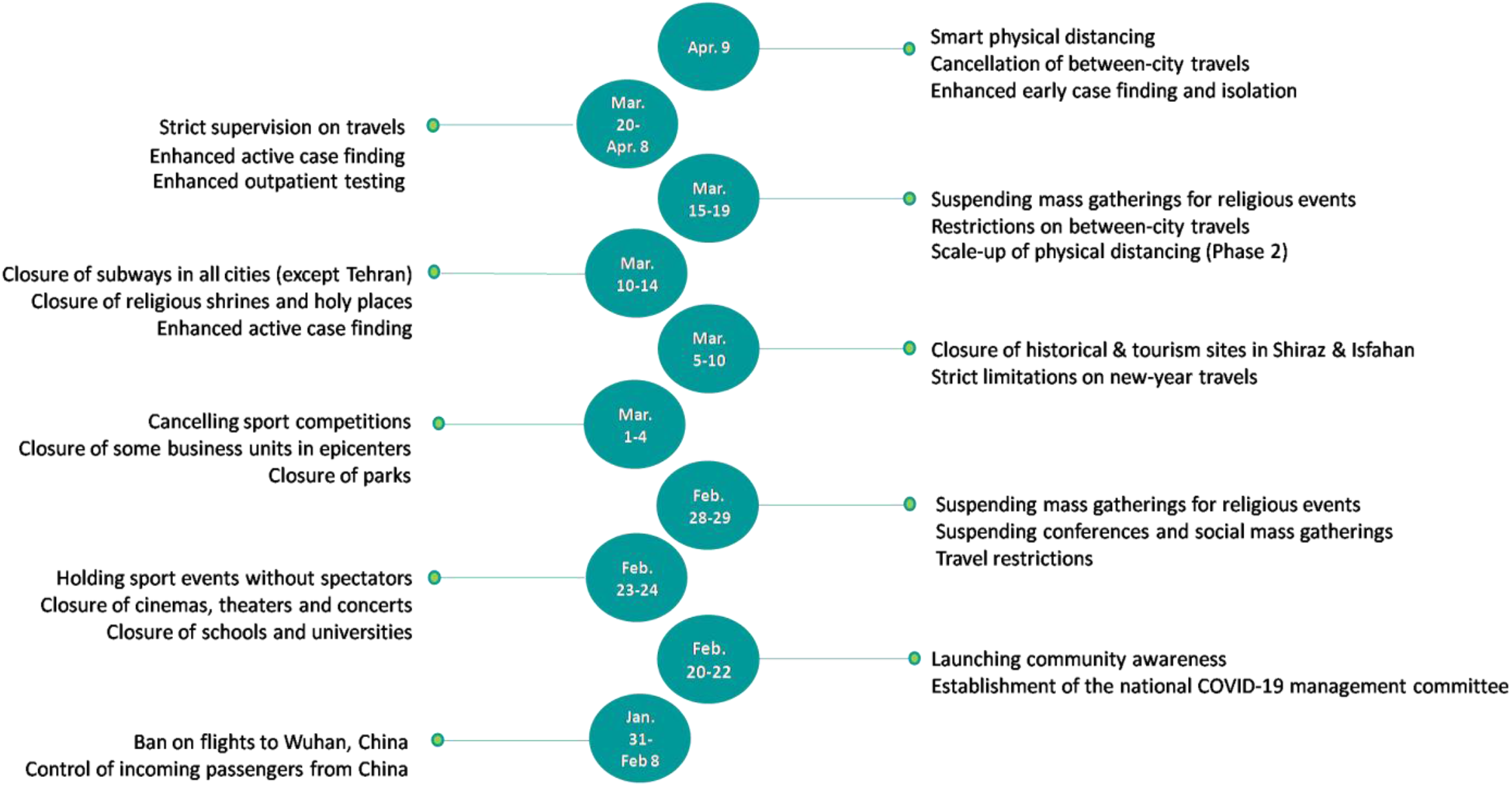
Iran’s interventions aimed at controlling the COVID-19 epidemic since beginning of the outbreak.

## Methods

### Model description

We formed a compartmental model to estimate the total number of COVID-19 patients, hospitalizations and deaths in Iran and its capital city of Tehran (Figure 2). We used an extended Susceptible-Exposed-Infected/Infectious-Recovered/Removed (SEIR) model that divides the target populations (i.e., Iran and Tehran as the capital) into different compartments. The conceptual framework of the COVID-19 transmission model is presented in Appendix AA. In brief, we considered the following comportments: a) *susceptible*, referring to the total number of individuals (i.e., hosts) who have been susceptible to COVID-19. We assumed the entire population as susceptible in our model; b) *exposed*, referring to individuals who are exposed to COVID-19 while they are asymptomatic and not yet infectious; c) *infected*, referring to infected people who demonstrate clinical symptoms after their incubation period and have the potential to transmit the disease to other susceptible individuals; and d) *recovered/removed*, depending on the severity of the disease. We assumed that the infected people will i) be recovered and immune from re-infection and therefore, no longer transmit the infection, or ii) have mild to moderate clinical symptoms while they follow home-isolation guidelines without requiring hospitalization, iii) have severe clinical symptoms and require hospitalization. These individuals would either be recovered (and then discharged) or fail to respond to treatment and pass away (removed) from the model. Monte Carlo method was used to build the 95% uncertainty intervals (UI) around the point estimates of the total expected numbers. To do this, we used the statistical distribution of a set of parameters obtained from both the existing evidence and expert opinion (listed in Table 1 Appendix BB). Data were analyzed using Vensim DSS 6.4E software.

**Table 1.** Different isolation scenarios for estimation of COVID-19-related infections, hospitalization, and deaths in Iran

| **Isolation Scenarios** | **Description** |
| --- | --- |
| **Scenario A** | Isolation for the entire period of the epidemic was considered to be 0% (the worst scenario) |
| **Scenario B** | Policy interventions were at the level of encouragement and the overall mean of isolation for the entire period of the epidemic was considered to be 10% |
| **Scenario C** | Isolation was considered to be 10% from January 21 to February 19, 15% after the initiation of the epidemic from February 20 to March 10, and finally 20% from March 11 to June 19, 2020, which are the results of the *minimal* possible interventions of the health system, behavior change of the public, and containment strategies |
| **Scenario D** | Isolation was considered to be 10% from January 21 to February 19, 15% after the initiation of the epidemic from February 20 to March 10, and finally 30% from March 11 to June 19, 2020, which are the results of the *moderate* possible interventions of the enhanced health system, social and behavioral change of the public (e.g., social distancing, hand washing), and containment strategies (e.g., closing schools and universities) |
| **Scenario E** | Isolation was considered to be 10% from January 21 to February 19, 15% after the initiation of the epidemic from February 20 to March 10, and finally 40% from March 11 to June 19, 2020, which are the results of the *maximum* possible interventions of the health system, behavior change of the public, and containment strategies. |

**Figure 2.**
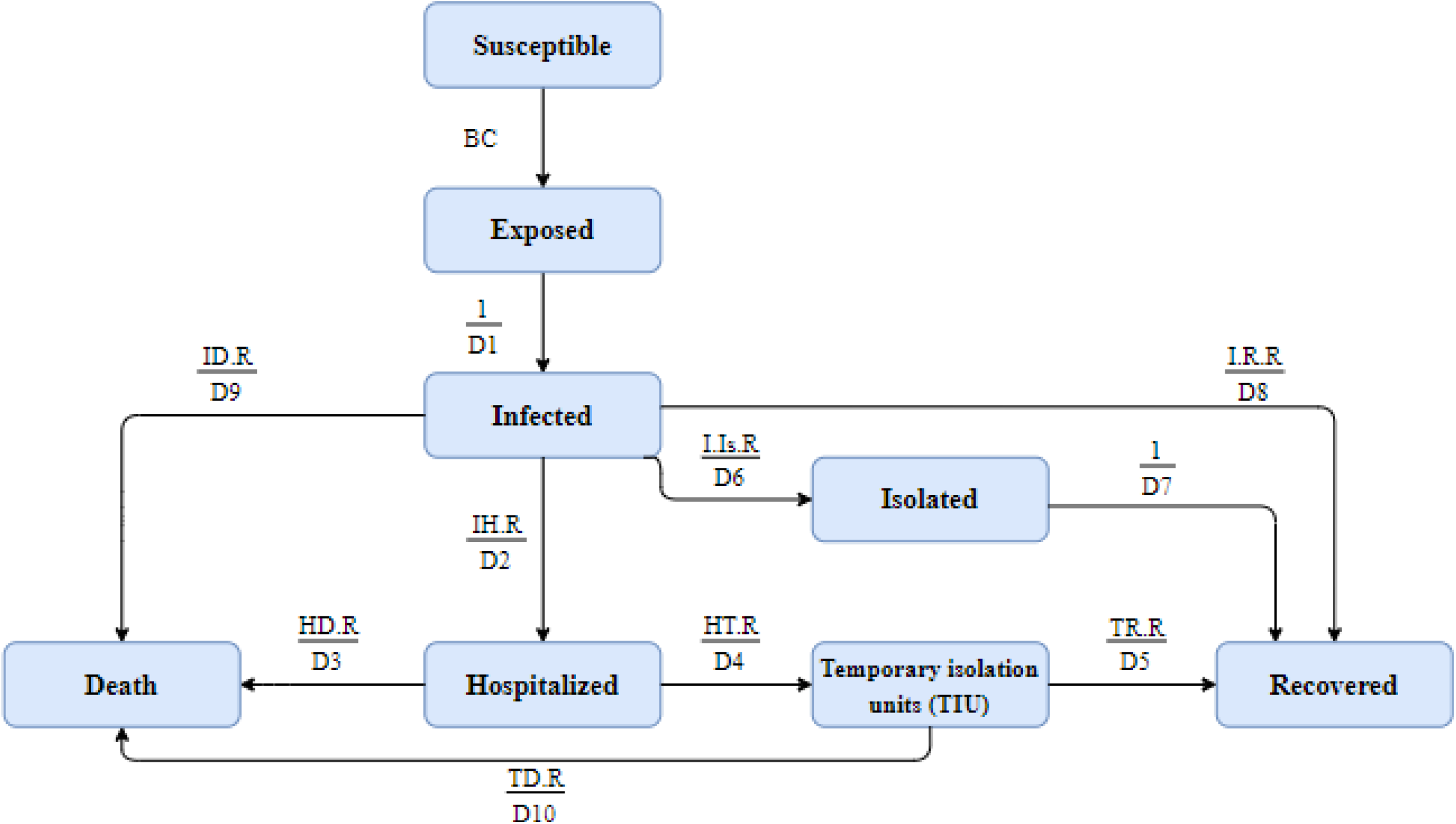
The SEIR conceptual model from susceptible to recovery or deaths.

### Model parameters and calibrations

Based on the country’s official reports and available epidemiological data, January 21, 2020 was considered as the initial day of the COVID-19 outbreak in Iran. We used several parameters as inputs for the model and obtained their values from a comprehensive literature review and published articles in relation to COVID-19, as well as some corresponding parameters and values considered for the similar epidemics, in particular, H1N1 influenza.^10^ We first shared the initial values of the parameters with Iran’s national and scientific committees and experts, and then made the necessary adjustments. We compared the revised values of these parameters with the literature as well as the pattern of the epidemic in Iran. We then made the final revisions for the values of the parameters used as input for the predicted number of COVID-19 infected cases and its associated deaths using the extended SEIR model. Input parameters and their brief descriptions are presented in Tables 1 and 2 in Appendix BB.

**Table 2.** The estimated date for epidemic peak, number of infected, hospitalized, and deaths in Iran and Tehran under five different scenarios from 21 January to 19 June, 2020.

|  | Scenario A |  | Scenario B |  | Scenario C |  | Scenario D |  | Scenario E |  |
| --- | --- | --- | --- | --- | --- | --- | --- | --- | --- | --- |
|  | Iran | Tehran | Iran | Tehran | Iran | Tehran | Iran | Tehran | Iran | Tehran |
| Infected cases |  |  |  |  |  |  |  |  |  |  |
| Peak date | 9-Jun | 19-Jun | 30-Apr | 30-May | 31-Mar | 20-Apr | 1-Mar | 31-Mar | 1-Mar | 11-Mar |
| Number (95% UI*) of new cases per day on peak | 66,000<br>(15,600, 145,000) | 19,000<br>(5,000, 36,000) | 16,500<br>(5,400, 35,000) | 4,600<br>(1,000, 11,000) | 10,700<br>(4,600, 20,000) | 2,200<br>(700, 4,700) | 9,400<br>(5,600, 15,000) | 1,700<br>(700, 3,300) | 9,400<br>(5,600, 15,000) | 1,600<br>(900, 2,600) |
| Total number (95% UI*) until 19 June 2020 | 5,196,000<br>(1,753,000, 10,220,000) | 1,228,000<br>(416,400, 2,407,000) | 1,643,300<br>(603,400, 3,380,000) | 398,600<br>(131,000, 852,000) | 949,600<br>(368,000, 1,960,000) | 211,800<br>(71,700, 467,700) | 689,000<br>(286,000, 1,363,000) | 142,000<br>(51,200, 309,800) | 550,000<br>(243,500, 1,070,200) | 106,400<br>(41,200, 225,000) |
| Hospitalized cases |  |  |  |  |  |  |  |  |  |  |
| Peak date | 19-Jun | 19-Jun | 12-May | 10-Jun | 14-Apr | 28-Apr | 15-Mar | 10-Apr | 14-Mar | 15-Mar |
| Number (95% UI*) of the existing cases on peak | 67,800<br>(25,100, 131,000) | 19,000<br>(7,800, 34,200) | 14,600<br>(5,800, 28,100) | 4,000<br>(1,200, 8,600) | 7,800<br>(3,200, 14,500) | 1,600<br>(550, 3,400) | 6,600<br>(3,500, 10,900) | 1,100<br>(400, 2,100) | 6,500<br>(3,400, 10,600) | 1,000<br>(500, 1,800) |
| Total number (95% UI*) until 19 June | 966,000<br>(467,800, 1,702,000) | 228,000<br>(108,800, 400,800) | 279,500<br>(119,200, 527,700) | 65,500<br>(26,100, 127,700) | 147,000<br>(56,200, 292,300) | 32,000<br>(11,000, 67,700) | 98,700<br>(38,100, 195,600) | 20,000<br>(6,700, 42,700) | 74,500<br>(29,400, 146,500) | 14,000<br>(4,900, 29,700) |
| Deaths |  |  |  |  |  |  |  |  |  |  |
| Total number (95% UI*) until 19 June | 111,000<br>(53,400, 200,000) | 25,800<br>(12,000, 47,000) | 32,900<br>(14,700, 62,000) | 7,600<br>(3,100, 15,000) | 17,700<br>(7,300, 34,400) | 3,800<br>(1,400, 7,900) | 11,000<br>(4,500, 21,700) | 2,200<br>(800, 4,600) | 7,800<br>(3,200, 15,000) | 1,500<br>(500, 3,000) |
| Total Case fatality rate | 2.2% | 2.1% | 2.0% | 1.9% | 1.9% | 1.8% | 1.6% | 1.5% | 1.4% | 1.3% |
| * UI: Uncertainty Intervals |  |  |  |  |  |  |  |  |  |  |

The impact of seasonality (a sinuses’ function) was considered in calculating the transmission probability (beta coefficient) of the disease, indicating the potential for some level of change in transmissibility of the virus from one season to another.(10) Therefore, we assumed that COVID-19 might behave the same as influenza such that the transmission of the virus may tend to reduce by approaching to warm seasons (i.e., spring and summer). We then considered the end of December in winter with the most transmission probability and the end of June in summer with the least transmission probability. The minimum and maximum values of the seasonal changes were considered to be 0.02 and 0.045, respectively. A time-varying state was considered for the effective contact rate (C parameter). We first incorporated value of 14 in the Tehran model and 13 in the national model in the early weeks of the epidemic.^11-15^ After the announcement of the epidemic by the officials, multiple public health measures implemented as a response to the epidemic to reduce contact rates and then transmission rates in public. Approaching the assumed end of the epidemic, we considered value of 5 for this parameter, with some fluctuations due to Nowruz holidays within this period (Table 2 in Appendix BB). Five possible scenarios were considered for isolation of the infected cases (Table 1).

## Results

### Expected number of infected cases (Table 2, Figure 3)

#### Iran

Under scenario A, the epidemic peaks on June 9 at around 66,000 (95% UI: 15,600-145,000) new infected cases per day. Total number of infected cases by June 19 is expected to be 5,196,000 (95% UI: 1,753,000-10,220,000). Under scenario B, the peak occurs on April 30 at around 16,500 (95% UI: 5,400-35,000) infected cases a day; a total of 1,643,300 (95% UI: 603,400-3,380,000) infections occurred by June 19. Under scenario C, the peak occurs on March 31 at around 10,700 (95% UI: 4,600-20,000) infected cases a day; a total of 949,600 (95% UI: 368,000-1,960,000) infections occurred by June 19. The number of infected cases at the peak time under scenario D (9,400 cases on March 1) and scenario E (9,400 cases on March 1) are further decreased. The total number of infected cases by June 19 under scenario D (689,000 cases on March 1) and scenario E (550,000 cases on March 1) are further decreased.

**Figure 3.**
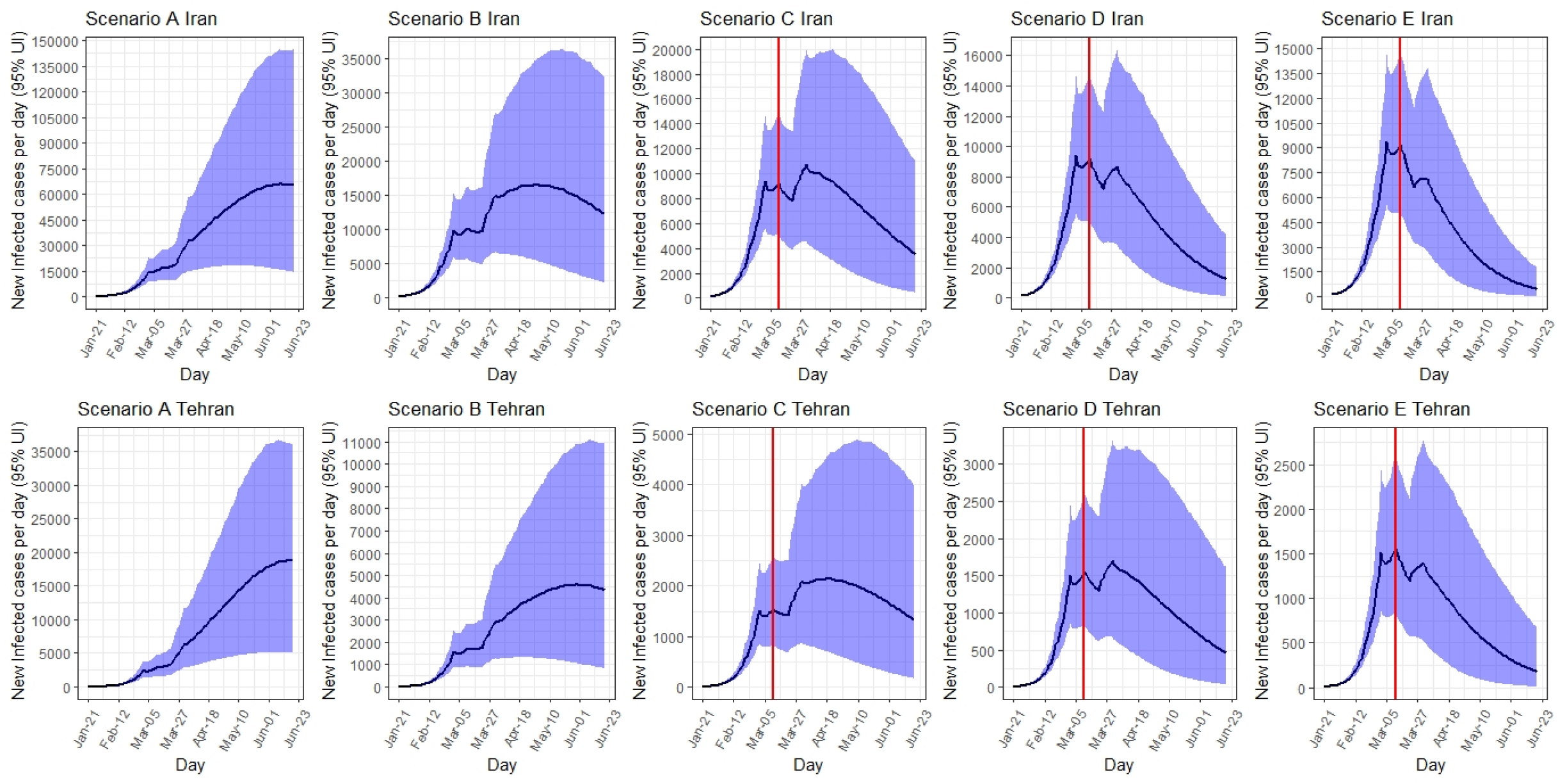
The estimated number of new infected cases per day in Iran and Tehran under five different scenarios from 21 January to 19 June, 2020. [*the red line shows the time when intervention was started].

#### Tehran

Under scenario A, the number of new cases per day in Tehran peaks on June 19 at around 19,000 (95% UI: 5,000-36,000). Total number of infected cases by June 19 is expected to be 1,228,000 (95% UI: 416,400 to 2,407,000). The number of new cases at peak and total number of cases decreased to lowest number from scenario B to E. Under scenario E, the number of new cases per day would reduce to 1,600 (95% UI: 900-2,600), and the total number of infected cases by June 19 would reduce to 106,400 (95% UI: 41,200-225,000).

### Expected number of hospitalized cases (Table 2, Figure 4)

#### Iran

Under scenario A, the number of hospitalized cases peaks on June 19 at around 67,800 (95% UI: 25,100-131,000) per day. Total number of hospitalized cases by June 19 is expected to be 966,000 (95% UI: 467,800-1,702,000). Under scenario B, the peak occurs on May 12 at around 14,600 hospitalized cases a day. Under scenario C, the peak occurs on April 14 at around 7,800 hospitalized cases a day. The number of hospitalized cases at the peak time under scenario D was 6,600 cases (on March 15) and under scenario E was 6,500 cases (on March 14).

**Figure 4.**
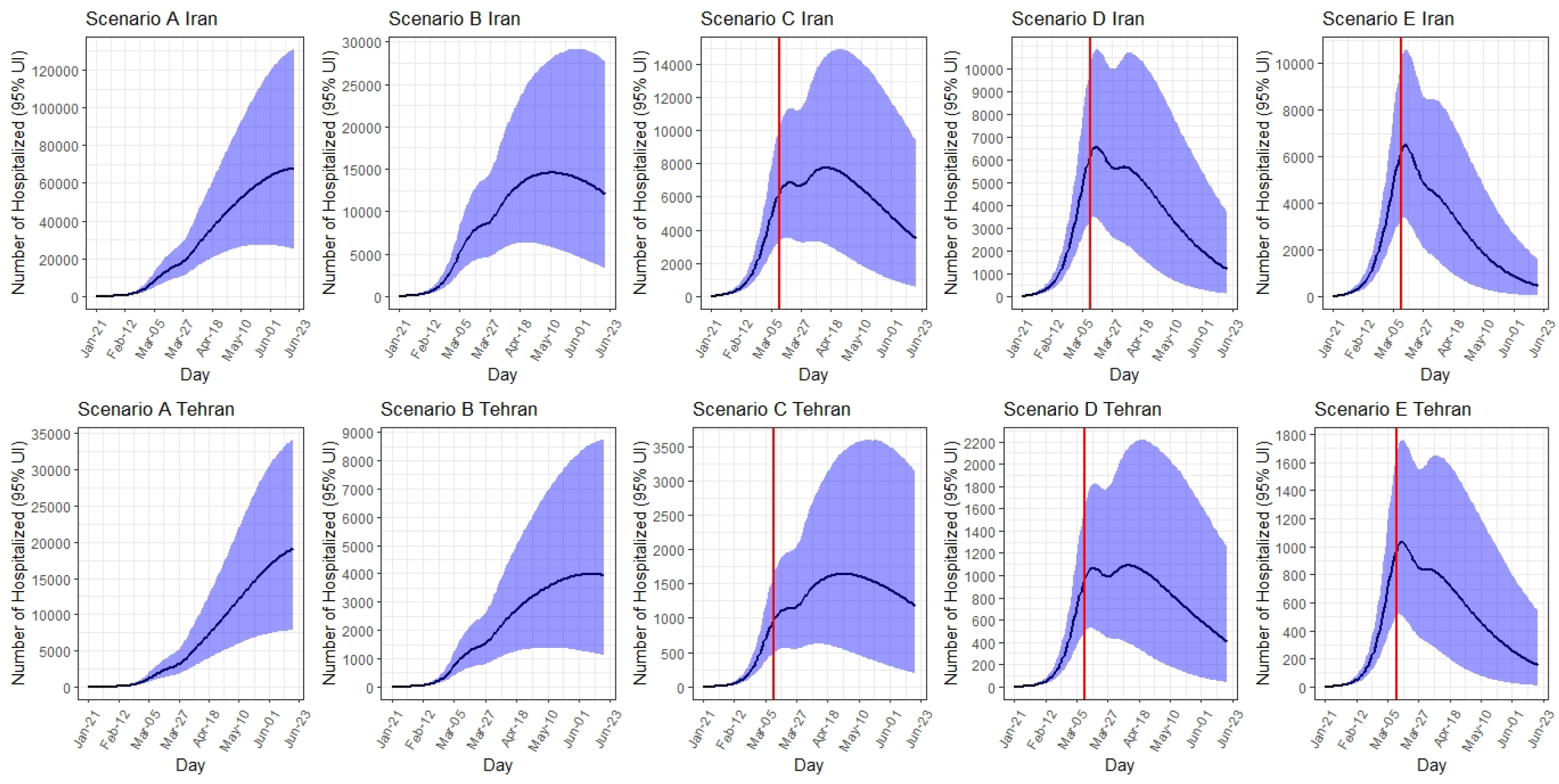
The estimated number of existing hospitalized cases in Iran and Tehran under five different scenarios from 21 January to 19 June, 2020. [*the red line shows the time when intervention was started].

#### Tehran

Under scenario A, the number of hospitalized cases per day in Tehran peaks on June 19 at around 19,000 (95% UI: 7,800-34,200). Total number of hospitalized cases by June 19 is expected to be 228,000 (95% UI: 108,800 – 400,800). The number of hospitalized cases at peak and total number of hospitalized cases are decreased to lowest number from scenario B to E. Under scenario E, the number of hospitalized cases per day decreases to 1,000 (95% UI: 500 - 1,800), and the total number of hospitalized cases by June 19 decreases to 14,000 (95% UI: 4,900-29,700). **Expected number of deaths (Table 2, Figure 5)**

**Figure 5.**
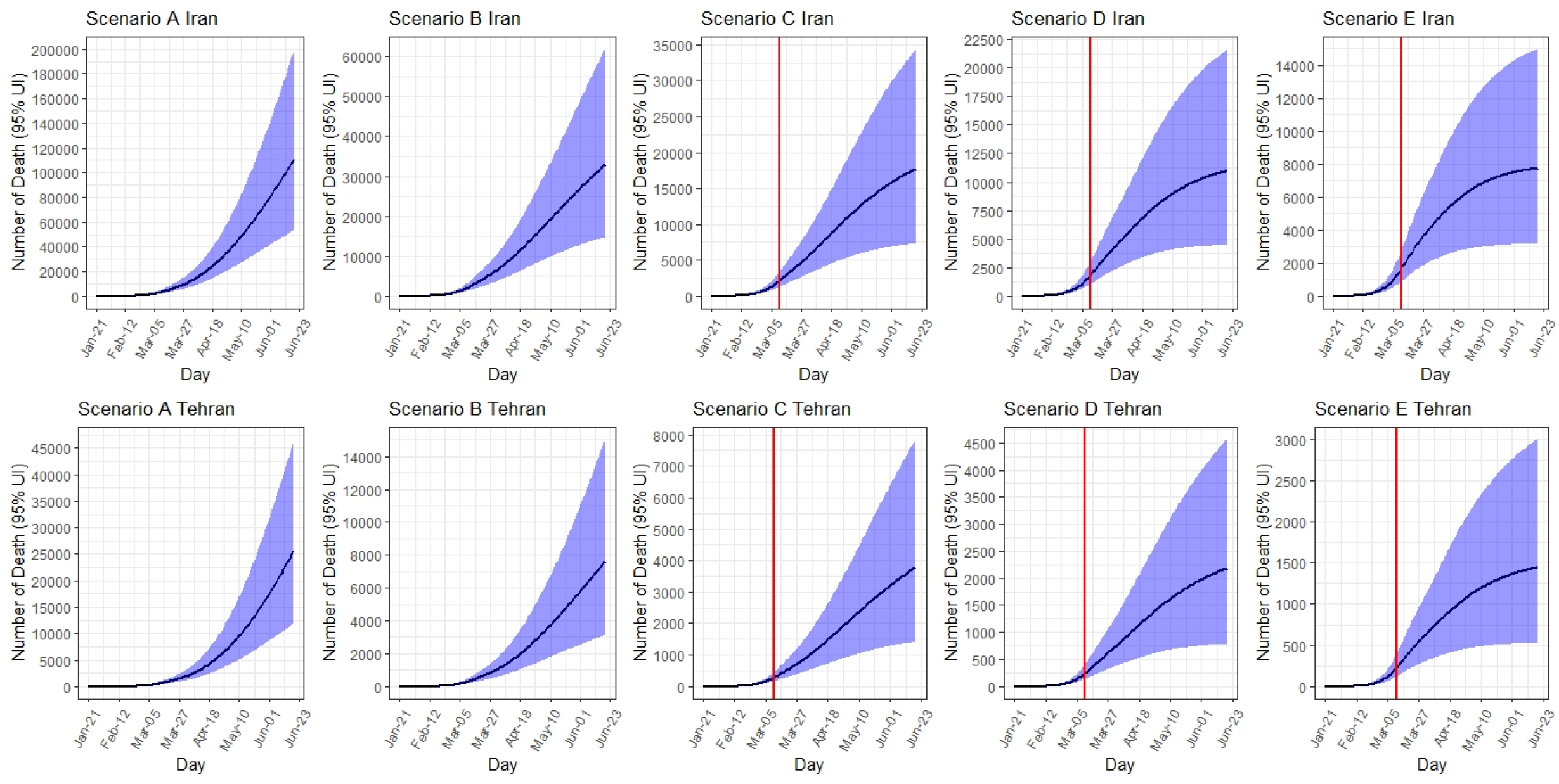
The estimated number of deaths in Iran and Tehran under five different scenarios from 21 January to 19 June, 2020. [*the red line shows the time when intervention was started].

#### Iran

Up to June 19, the total expected number of deaths ranges from 111,000 (95% UI: 53,400 to 200,000) under scenario A to 7,800 (95% UI: 3,200-15,000) under scenario E. The corresponding case fatality rate under scenario A was 2.2%, which would decrease to 1.4% under scenario E.

#### Tehran

Up to June 19, the total expected number of deaths ranges from 25,800 (95% UI: 12,000 to 47,000) under scenario A to 1,500 (95% UI: 500 to 3,000) under scenario E. The corresponding case fatality rate under scenario A was 2.1%, which would decrease to 1.3% under scenario E.

## Discussion

Our modeling exercise showed that with no intervention (i.e., scenario A), more than five million COVID-19 infections would occur in Iran till mid-June, of whom 18.9% would be hospitalized and 2.1% would die. However, under the best-case scenario (i.e., scenario E), the number of infected cases could be reduced by 90%, hospitalizations by 92%, and deaths by 93%. Our projection in scenario C, which is a middle ground scenario, appeared to be aligned with the current statistics from the national reports. Even under scenario C, the burden of the epidemic in Iran will be large, last for several months and might surpass the current capacity of the healthcare system. Our model indicated that under scenario B, Iran might have sufficient hospital bed capacity for COVID-19. As of April 8, 2020, public hospitals in Iran, which are mainly responsible for the COVID-19 response, had 146,137 beds (9,134 in intensive care units [ICU] and 9,730 ventilators). By discharging inpatient cases and postponing elective medical care and surgeries, Iran has currently allocated 100,437 empty beds, 5,790 empty ICU beds, and 4,650 ventilators towards COVID-19 patients. This might be sufficient for sever COVID-19 patients as predicted by scenario B which would still require appropriate supplies and staffing. As of April 8, 2020, there were 0.41 physician and 1.14 nurse per public hospital bed in Iran. Even under scenario C, the shortage of healthcare workers would be challenging.

In addition to individual- and community-level physical distancing control measures that have been implemented in Iran from late February to late March, the number of testing for COVID-19 screening was also increased from 1,000 tests per day on March 12, 2020 to about 10,000 tests per day on April 11, 2020. This has led to an increase in case finding, contact tracing of new cases, and isolation of confirmed cases. Despite the persistent increase in testing, the number of confirmed cases per day peaked at 3,200 on March 30 and then decreased to 1,374 cases per day on April 18.^16^ Moreover, measuring the real intensity and coverage of physical distancing in Iran is challenging as these practices have not been closely followed during to the new year (i.e., Nowruz) holidays between March 19 and 26.^17,18^ Such a significant increase in travels made the government to enforce physical distancing by closing the roads. While we did account for the Nowruz effect in our model by increasing the number of contacts per day (parameter c) from 5 to 6 from 21 to 31 March, the true effect of Nowruz is still unknown on the current and future projected numbers.

Based on the peak time for the number of confirmed cases per day, scenarios C is likely to be the most plausible one for COVID-19 outbreak in Iran. Under this scenario, 10,100 new infections occurred on April 5, which is four times the number of confirmed cases. This scenario expects the total number of infections and deaths by mid-June to be around 950,000 and 17,600, respectively; around four times the number of observed deaths to date (i.e., 5,118 cases by April 19). Our findings however, suggest that scenario B is the most plausible scenario in the Capital city of Tehran which highlights the importance of implementing special measures and policies in this city to reduce the spread of the disease. Our findings in scenario B are comparable with other dynamic model estimations in Iran,^19^ that have predicted around 1.6 million (90% UI, 0.9M – 2.6M) cases, and 58,000 (90% UI, 32,000-97,000) deaths under their most optimistic scenarios. However, this model had three structural assumptions on three major parameters (i.e., duration of illness as 14 days, asymptomatic period as 4 days, and transmission probability as 0.25 during the asymptomatic period) and did not account for seasonality distribution.^19^

Our projections of the COVID-19 morbidity and mortality in Iran under different physical distancing and isolation scenarios are insightful for government’s policies regarding relaxing the physical distancing and isolation interventions. Although the decision on the timing of and approaches towards lifting the physical distancing restrictions is extremely difficult and varies across countries with different economic and healthcare infrastructures, it is critical to follow an evidence-informed approach to avoid the second and further waves of COVID-19 epidemics in Iran. A modeling study from China for example,^15^ showed that a stepwise (25% of the workforce working in weeks 1 to 2, 50% of the workforce working in weeks 3 to 4, and 100% of the workforce working and school resuming from week 5 forward) return to work or school at the beginning of April (about five months after the first case reported from China), is much more effective than the beginning of March. This study estimated that just a one-month delay in the stepwise lifting of the physical distancing measures would reduce the magnitude (92% by mid-2020, 24% by end-2020) the epidemic and delay its peak by two months and therefore avoid overwhelming the healthcare systems.^15^

In Iran, a recent executive order from the government lifted the restrictions on nationwide business shutdown and allowed most people to return to work only 2.5 months after the identification of the first COVID-19 case in Iran.^9,20^ The government has also planned to reopen schools in low-risk cities and non-essential low or medium-risk jobs (e.g., all production units in industrial, business, technical service, and distributional sections), as well as removing the shelter-in-place order and resuming domestic and international travels. These decisions are mainly derived from the Iranian government’s economic challenges that have been elevated by the comprehensive sanctions imposed by the USA.^21,22^ While these concerns are understandable and longer shutdown of an already overstretched economy is a tough decision and would be very taxing on the government and the public, our findings as well as lessons learned from China,^15,23,24^ suggest that this approach is not justified by evidence and would most likely risk overwhelming the healthcare systems with soon-to-come second and further waves of COVID-19 epidemic in Iran; costs that might surpass the marginal economic benefits of opening businesses and schools a few months sooner. Nonetheless, it is fortunate that the Iranian CDC is now planning to shift from physical distancing to targeted case-finding, intensify contact tracing and careful isolation of identified cases as well as self-quarantine of symptomatic people.

Our study had three major limitations. First, some of the key parameters (e.g. hospitalization rate, incubation period, transmission probability) that were used in the model were from other countries or expert opinion, but not from empirical data from Iran. To address this limitation, we reported a range of uncertainty intervals. Second, the uncertainty intervals are fairly wide for most of our results which resulted from the uncertainty in model parameters. Third, our projection for the course of the epidemic in the coming months in Iran is based on the assumption we made about implementing and sustaining the physical distancing as planned, and any changes in policy and public interventions may change the course of the epidemic. Despite these limitations, our results under different scenarios provide a foundation to measure the effect of interventions that are ongoing in the country.

With no approved vaccination, prophylaxis or therapy, we found physical distancing and isolation that includes public awareness and case-finding/isolation of 40% of the infected cases could reduce the burden of COVID-19 in Iran by 90% by mid-June.

## Data Availability

All necessary data are reported.

## Declaration of interests

Except for the senior author, Dr. Ali Akbar Haghdoost, who is the Deputy Minister of Health and the Head of National COVID-19 Committee, the rest of the authors declare no conflict of interest, real or perceived.

## Funding

The authors received no funding for this work.

## Ethical approval

The proposal of the present study was approved by ethics committee of Kerman University of Medical Sciences, Kerman, Iran (reference 98001239).

## Appendix AA The Susceptible-Exposed-Infected/Infectious-Recovered/Removed (SEIR) model

### A. The SEIR conceptual model

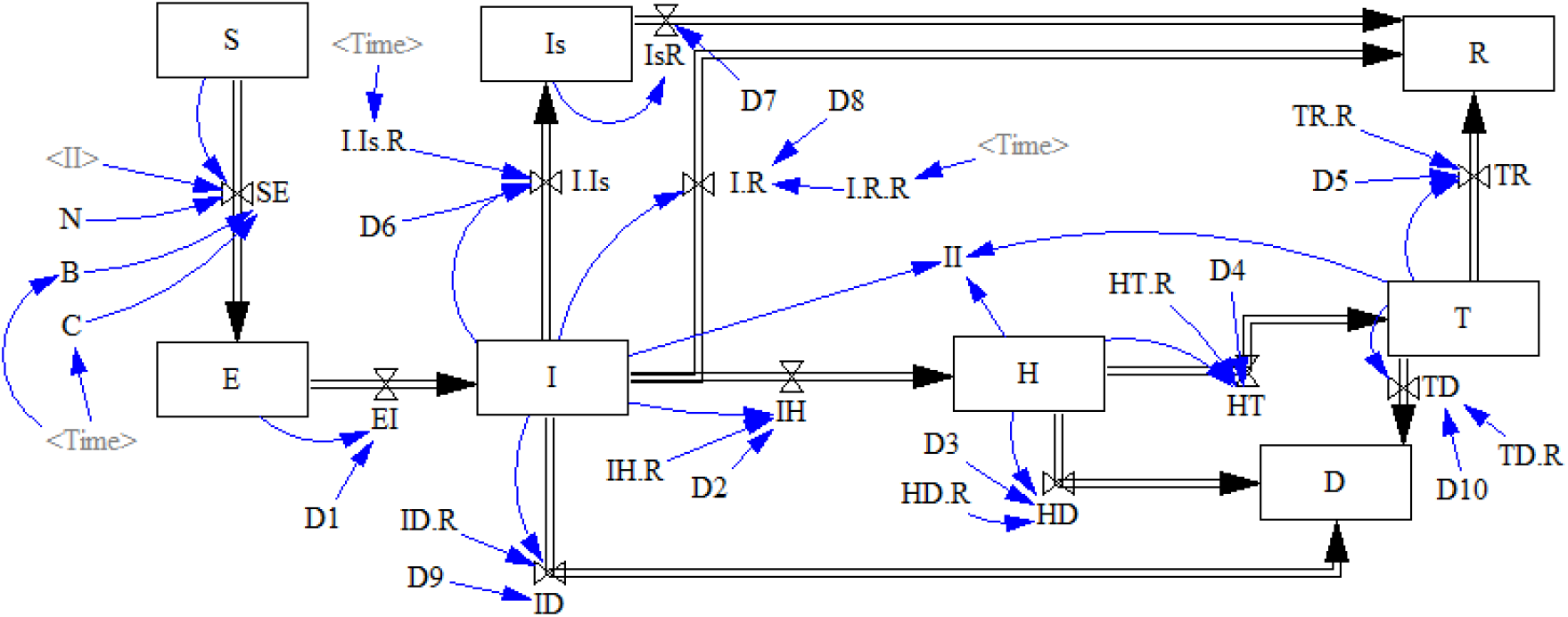

### B. Model formation

As mentioned in the main text, we used an extended form of the Susceptible-Exposed-Infected/Infectious-Recovered/Removed (SEIR) model, which is a dynamic modelling approach. Such epidemiological models are assumed to be compartmental in which the target population will be divided into different sections or compartments. The conceptual framework of our model is shown above in section A. Using this model, we divided the population of size of the target populations (mainland as the total country population and Tehran as the country capital) into the following comportments:

a. Individuals who are *susceptible* (S(t)): an individual (i.e., host) is initially assumed to be susceptible to the virus (i.e., COVID-19 here), and that the virus can be transmitted from infected individuals to susceptible individuals. In this model, the entire population of the target populations were considered to be susceptible. The differential equation of this compartment is shown in Equation 1:

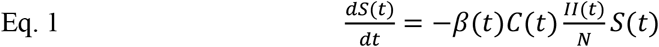

where β(t) indicates transmissibility of the virus, and C(t) indicates the contact rates, and II(t) refers to the total number of infected people who transmit the infection, calculated as Infect (t)+(0.1×Temporary Isolation Units)+(0.02×Hospital) (explained below).
b. Individuals who are *exposed* (E(t)): refers to individuals who are exposed to the infection, but they are not yet infectious. These individuals are asymptomatic in this period. The differential equation of this compartment is shown in Equation 2:

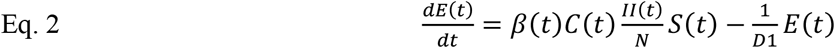
c. Those who are *infected* (I(t)): in this model, infected patients after a period of time will demonstrate clinical symptoms of the infection and will transmit the infection to any other susceptible individuals. Equation 3 shows the differential equation of this compartment:

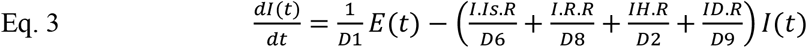
d. Those who *recovered*: depending on the severity of the infection, infected individuals will end up with one of the following four states:
  i. Infected individuals who are recovered (R(t)), who will be assumed to be immune from re-infection and no longer transmit the infection (recovered box in Fig 1). Equation 4 shows the differential equation of this section:

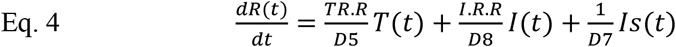
  ii. Infected individuals who will have mild to moderate clinical symptoms, but they will be home-isolated without requiring hospitalization (IS(t)), and they will be recovered (isolated box in Fig 1). Equation 5 shows the differential equation of this section:

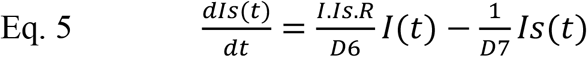
  iii. Infected individuals who will have severe clinical symptoms requiring hospitalization (hospitalized box in Fig 1). These individuals will have two possible outcomes: i) some hospitalized cases will be recovered and then discharged (T box in Fig 1), or ii) some will not respond to the medical care and die (death box in Fig 1). Equations 6 and 7 show the differential equation of hospital box (H(t)) and T box (T(t)) respectively:

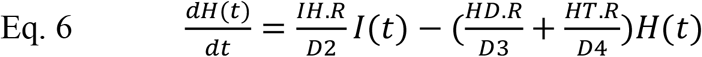

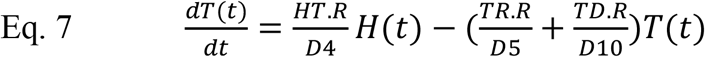
  iv. Infected individuals who will die (removed) without recovery (D(t)) (move to death box). Equation 8 shows the differential equation of this section:

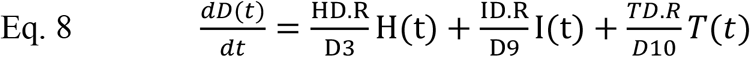

## Appendix BB

**Table 1.**
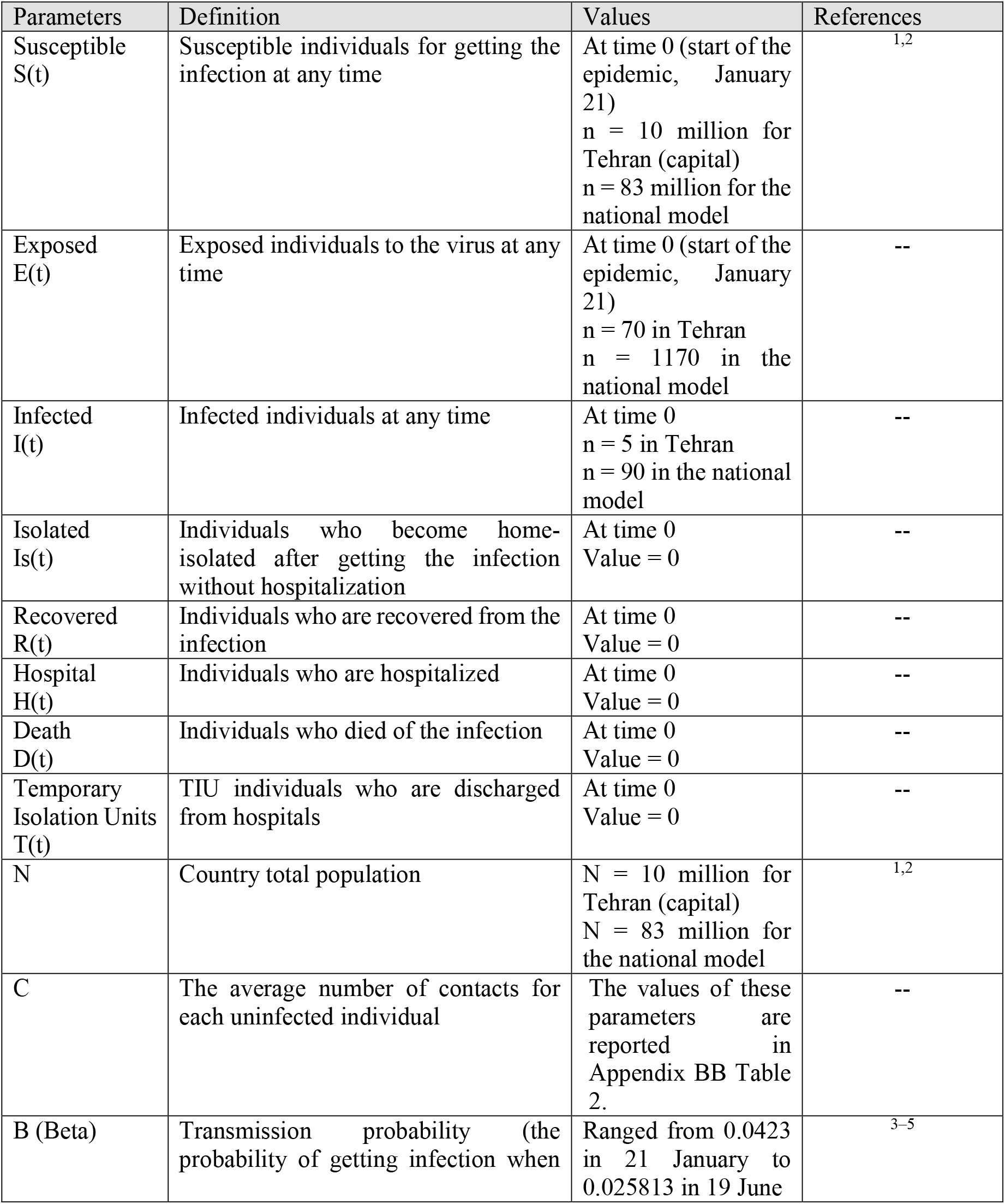
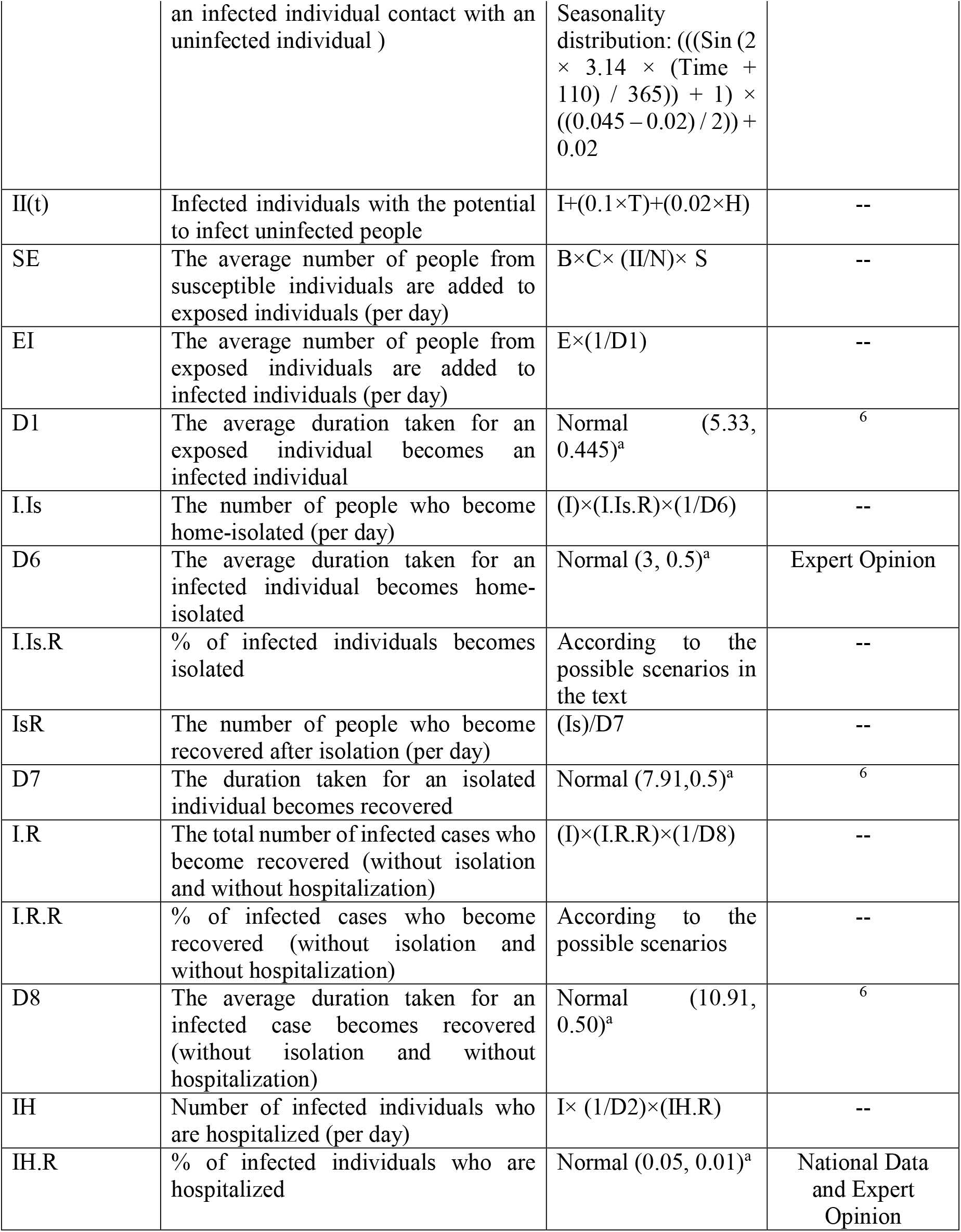
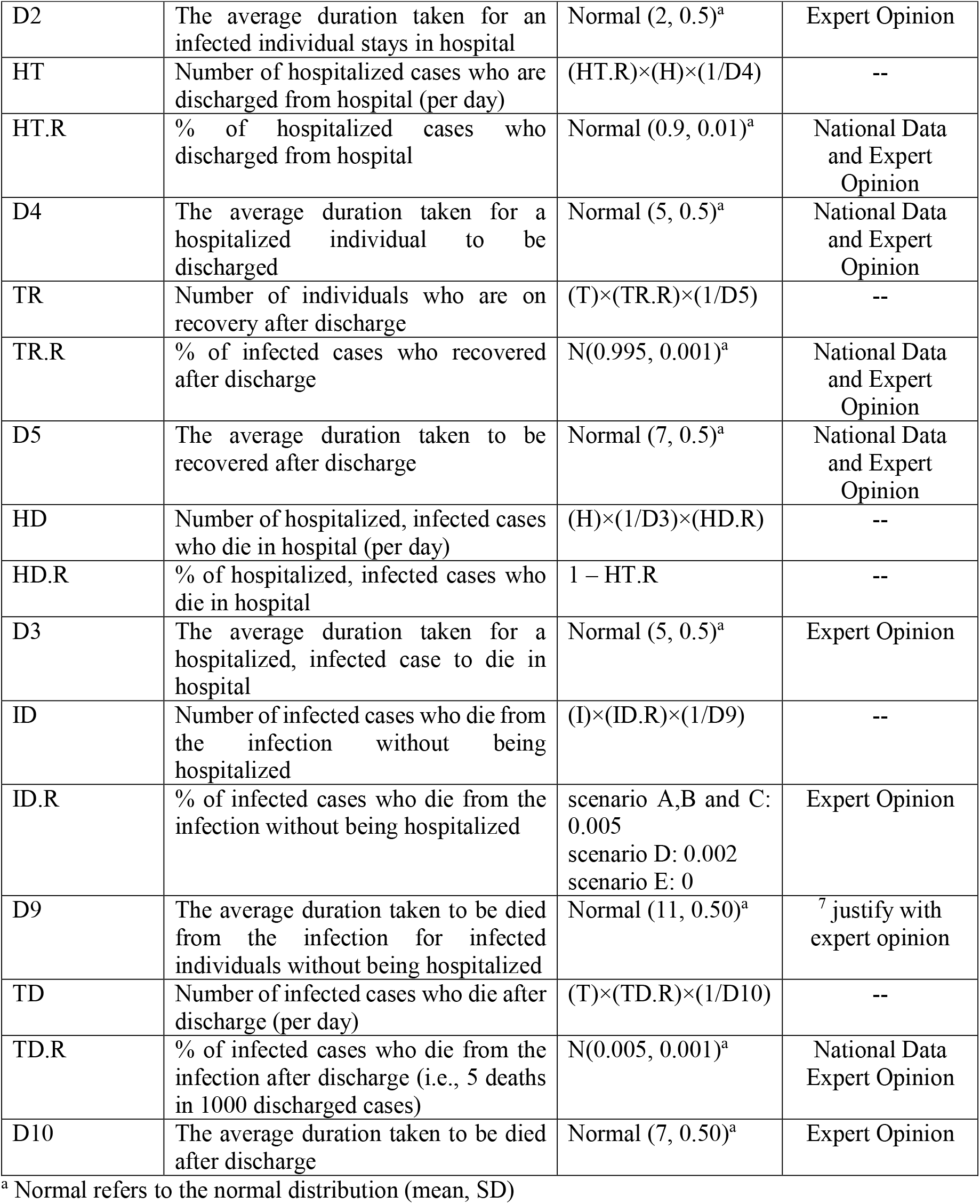
Disease and demographic inputs and parameters used in the SEIR model

**Table 2.** The values considered for effective contact rate (i..e, C) within four months of the COVID-19 epidemic stratified by Tehran and national models

| Months and 10-day intervals | Contact rate (C) |  | Reference |
| --- | --- | --- | --- |
|  | Tehran model | National model |  |
| From Jan 21, 2020, to Jan 30, 2020 | 14 | 13 | 7–11 |
| From Jan 31, 2020, to Feb 9, 2020 | 13 | 12 |  |
| From Feb 10, 2020, to Feb 19, 2020 | 12 | 11 |  |
| From Feb 20, 2020, to Feb 29, 2020 | 10 | 9 |  |
| From Mar 1, 2020, to Mar 20, 2020 | 5 | 5 |  |
| From Mar 21, 2020, to Mar 31, 2020 | 6* | 6* |  |
| From Apr 1, 2020, to June 19, 2020 | 5 | 5 |  |

\* Contact rates were assumed to increase due to the Nowruz holidays within these periods

## Notes

### Competing Interest Statement

The authors have declared no competing interest.

### Funding Statement

No funding was received for this work.

